# Geographic and Phylodynamic Distribution of SARS-CoV-2 from Environmental Origin

**DOI:** 10.1101/2021.09.13.21263432

**Authors:** Elijah Kolawole Oladipo, Emmanuel Oluwatobi Dairo, Ayodele Eugene Ayeni, Boluwatife Ayobami Irewolede, Moyosoluwa Precious Oyewole, Charles Oluwaseun Adetunji, Julius Kola Oloke

## Abstract

The coronavirus disease 2019 (COVID-19) caused by the severe acute respiratory syndrome coronavirus 2 (SARS-CoV-2) has spread globally. Understanding the transmission dynamics of SARS-CoV-2 contamination in the environment is essential for infection control policies. This study aims to provide a phylodynamic analysis and distribution pattern of SARS-CoV-2 from the environment in terms of Source, clades, lineages, and their location. Ninety (90) retrieved whole-genome sequences of environmental sources from GISAID were investigated to determine the evolutionary process of SARS-CoV-2 and mutation in the isolated nucleotide sequences. The analysis was carried out using R, MAFFT, and MEGA X software. Out of the five countries studied, Austria has the highest distribution with sixty-five samples (72.2%), and the highest isolates of 68 (75.6%) were from raw sewage. The highest clade in circulation as obtained from the study is G with lineages B. The phylogeny of SARS-CoV-2 whole-genome sequences from Austria, the United States, China, Brazil, and Liechtenstein indicated that the SARS-CoV-2 viruses were all clustered together, irrespective of sequence geographic location. The study concluded by demonstrating a clear interconnection between the phylogeny of SARS-CoV-2 isolates from various geographic locations, all of which were locked in the same cluster regardless of their environment specimen. Thus, depicting the possibility of their origination from a common ancestor.

**Highlight:**

- Environmental sources of specimen isolated from raw sewage have the highest occurring specimen sequence, while those from breathing air, door handle, and wastewater have the lowest sequence.
- Environmental surveillance of SARS-CoV-2 is of great importance to control the spread of the virus. Untreated raw sewage should be of more priority for the environmental surveillance of the virus.
- Eighteen (18) nucleotide sequences from this study’s multiple sequence alignment shared a 90% similarity with the Wuhan-Hu-1 reference genome, indicating a common evolutionary origin.

## INTRODUCTION

Coronavirus SARS-CoV-2 is a novel RNA virus that belongs to a group of viruses that include SARS and MERS. There are more than 37 coronaviruses in the Coronaviridae family. Within the order, Nidovirales are responsible for causing COVID-19 which is responsible for causing disease that has an overwhelming effect on health care systems, economies, and societies of affected countries (Chin et al., 2020). Its outbreak was probably linked to a seafood market in the Wuhan City of China, where there is no credible evidence to support the rumors (Giovanetti et al., 2020; Liu et al., 2020; Paraskevis et al., 2020). Still, investigations from evolutionary, genomic, and proteomic studies suggested that bats could be the natural host of the novel virus (Liu et al., 2020). Also, a study conducted by Liu et al. points out that the SARS CoV-2 virus has a 96.2% sequence resemblance with bat CoV, RaTG13, and also shares 79.5% similarity with SARS CoV, given its name according to the Coronavirus Study Group of the International Committee on February 11, 2020. Structurally, they are positive-sense single-stranded RNA (ssRNA) virions with characteristic spikes projecting from the surface of the capsid protein (Neuman et al., 2006; Barcena et al., 2009). They also possess a genome of 30 kb in length, the largest among members of the RNA viruses, alongside a 5’ cap and a 3’ poly (A) tail, to enable their translation (Chen et al., 2020). Coronaviruses are made up of four main proteins, spike (S), membrane (M), envelope (E), and nucleocapsid (N) proteins. The spike protein (∼150 kDa) mediates its attachment to host receptor proteins, allowing their entrance into the cell (Collins et al., 1982). The membrane protein (∼25 to 30 kDa) attaches with nucleocapsid and maintains the curvature of the virus membrane (Neuman et al., 2011). As an integral membrane protein with ion channel activity, the envelope protein (8 to 12 kDa) is responsible for the virus’s pathogenesis by facilitating the assembly and release of virion particles (Ruch and Machamer, 2012). Nucleocapsid, the fourth protein, helps in packaging virus particles into capsids and promotes the formation of the replicase-transcriptase complex (RTC) (McBride et al., 2014).

The recent rise in the case of COVID-19 that started around December 2019 had raised significant concerns for many researchers across the globe to find lasting solutions to mitigate its effect. The ease of transmission of Coronavirus infections among diverse hosts is likely to be associated with its’ rate of mutation and high occurrence of recombination inherent in its genome (Neuman et al., 2006; Barcena et al., 2009).

SARS-CoV-2, a variant of the Coronaviridae family of viruses, which comprises viruses that cause respiratory and intestinal infections, spreads primarily via microdroplets, indicating its ability to survive in humid environments (Chin et al., 2020).

The environmental roles and the potential risk of environmental factors on the transmission and prevalence of COVID-19 pandemics, such as climate change, fomites, water transfer, air, and food, have been highlighted in some recent studies (Eslami and Jalili, 2020; Qu et al., 202; Wigginton et al., 2020). In the survey carried out by Kampf et al. (2020), emphasis was laid on the preeminence of inanimate surface contact to contribute to the transmission of SARS-CoV-2. SARS-CoV-2 tends to survive on a surface for days at room temperature, thus increasing its chance of transmission via touch (Kampf et al., 2020; Van et al., 2020). Moreover, some recent studies have also validated the presence of the SARS-CoV-2 virus in wastewater treatment facilities (Hindson, 2020; Medema et al., 2020; Naddeo and Haizhou, 2020; Wurtzer et al., 2020; Wu et al., 2020). As of 5 March 2021, 115,094,614 confirm cases of COVID-19 were recorded globally including, 2,560,995 deaths (WHO Coronavirus Disease situation Dashboard, 2021), in which an individual could likely have been infected through environmental Source. The fact that COVID-19 patients can spread the virus raises questions about SARS-CoV-2 infiltrating different ecological systems. SARS-CoV-2 had been detected in wastewater, cleaning swabs, air, sewage sludge, and river waters around the world (Patel et al., 2021).

The deaths and morbidities linked to the COVID-19 pandemic necessitate further investigation into the role of the ecosystem, such as wastewater and water bodies, in the disease’s transmission; based on the reviewed literature, the current wealth of information on coronaviruses in wastewater is minimal, necessitating further research (Patel et al., 2021). It is essential to track the emergence of SARS-CoV-2 diversity through comparative genomics, which may provide more knowledge about the virus’s prevention and transmission routes. Kampf et al. (2020) and Ong et al. (2020) reported that asymptomatic patients might increase the disease’s rapid dissemination by active shedding of the virus leading to direct transmission by droplet route and indirect transmission through contact with the contaminated environment. Even though the propagative numbers in SARS-CoV-1 (∼2.7) and SARS-CoV-2 are similar, they are lower than airborne measles, suggesting that air transmission is not the major transmission route (Guerra et al., 2017). The emphasis of this study was more on the environmental Source from which the sequences were isolated to provide clues on the most likely route the virus could be transmitted within a given geographical region. In this study, we carried out a detailed analysis on the phylodynamics of SARS-CoV-2 and its distribution in environmental sources to provide a thorough understanding of environmental contamination by SARS-CoV-2 to help broaden the scope of knowledge on viral transmission route and also facilitate proper infection control precautions.

## Materials and Methods

### Retriever of Data

Whole-genome sequences of SARS-CoV-2 isolates of environmental Source available in the GISAID database (https://www.epicov.org/epi3/frontend#57f16) as of 31 January 2021 was retrieved. A total of 101 SARS-CoV-2 complete sequences according to the filtering criteria (complete genome, environmental samples, high coverage level only, and low coverage excluded) were downloaded. The sequences came from 7 countries: Austria: 65 sequences, Brazil: 2 sequences, China: 16 sequences, Liechtenstein: 3 sequences, Hong Kong: 1 sequence, Netherlands: 6 sequences, and the USA: 8 sequences. Similarly, the sequences’ metadata was obtained from the GISAID database to obtain information about individual sequences. As a result of the missing information, which was inaccessible in the metadata collected online, 11 sequences were discarded out of the 101 sequences retrieved to arrive at 90 sequences. This, therefore, restricted the analyzed sequences to 5 countries as follows: Austria: 65 sequences, Brazil: 2 sequences, China: 14 sequences, Liechtenstein: 3 sequences, and the USA: 6 sequences. The table containing detailed information on the 90 SARS-CoV-2 sequences used in this study is provided as a Supplementary file (S1). The annotated reference genome sequence of the SARS-CoV-2 isolate Wuhan-Hu-1 (Accession number: NC_0455122) was downloaded from the National Center for Biotechnology Information (NCBI) GenBank database.

### Metadata Analysis

This study uses the mined data retrieved alongside the nucleotide sequences on the GISAID database to analyze the variability of the SARS-CoV-2 genome from the environmental Source, subsequently to the onset of the pandemic to ascertain its evolutionary relationship and geographical distribution. Metadata analyses were performed using R. Graphical distributions of SARS-CoV-2 were constructed based on their location, environmental Source, clade, and lineage. Moreover, a distribution graph was constructed on account of the Source, clades, and lineages in relation to the location where they are isolated. Emphasis focused on the environmental Source from which the sequences were found to provide clues on the possible routes by which this virus may be transmitted within a given geographical location. Understanding this will help implement appropriate ethical policies, guidelines, and procedures that could effectively handle various environmental materials to mitigate the exponential spread of COVID-19 and diminish the future strain on the environment.

### Multiple sequence alignment and evolutionary Analysis

The complete genome sequences of 90 SARS-CoV-2 obtained were aligned using MAFFT version 7 (https://mafft.cbrc.jp/alignment/server/) against the Wuhan-Hu-1 reference genome (Accession number: NC_0455122), with all parameters set at default (Katoh et al., 2019; Kuraku et al., 2013). Based on the whole genome sequences, a maximum likelihood tree was constructed using MEGA X with 50 bootstraps resampling to ascertain the common antecedent among each strain (Tingting et al., 2020).

## Result

### Metadata Analysis

The result obtained during this study indicated that 101 out of the sequences retrieved from GISAID, Ninety (90) genomic sequences were subjected to metadata analysis. The geographical distribution of the SARS-CoV-2 sequence based on their locations revealed that Austria has the highest distribution with sixty-five samples (72.2% of the obtained isolates) deposited into the database, followed by fourteen samples from China (15.6% of the isolates). Subsequently, a sum of six SARS-CoV-2 sequences from the United States of America (6.7%), while both 2 and 3 SARS-CoV-2 sequences from Brazil (2.2%) and Liechtenstein (3.3%) respectively, of the isolates as seen in Fig. 1.

**Figure 1:**
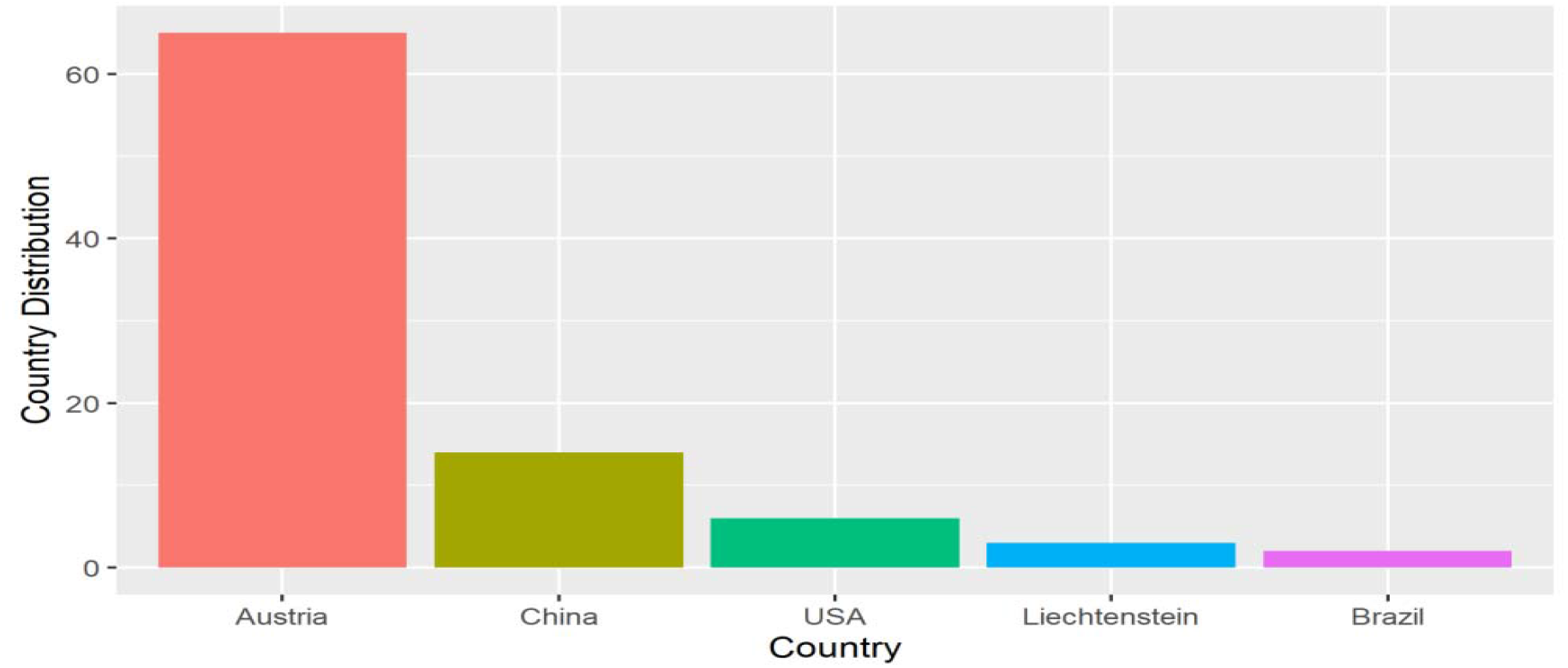
Country Distribution of SARS-CoV-2 from Environmental Source.

Moreover, based on environmental sources, specimen isolated from raw sewage has the highest distribution (68), occupying 75.6%. Also, nine (9) sequences were reported from the outer packaging of cold chain products, 10%, followed by six (6) sequences from an environmental swab of 6.7%. Both air and sewage have two sequences with 2.2% each while breathing air, door handle, and wastewater have one sequence with 1.1% respectively, as shown in Fig. 2.

**Figure 2:**
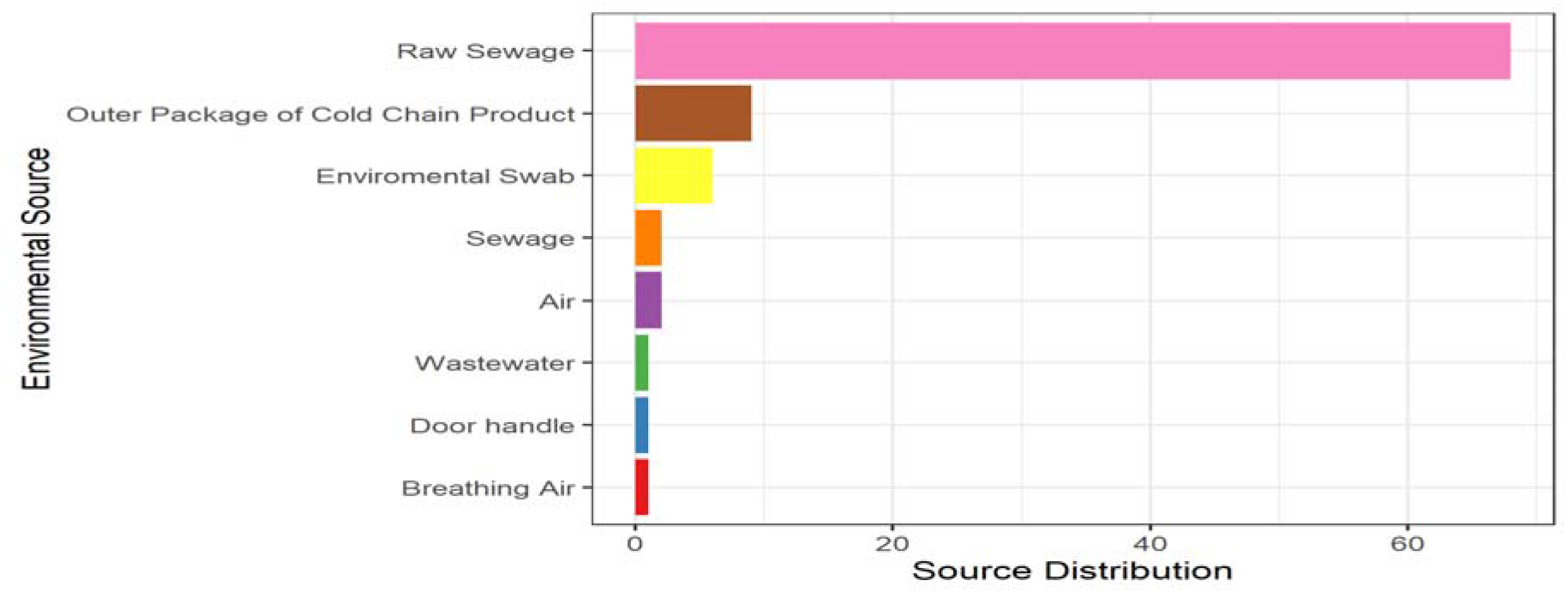
Environmental Source distribution of SARS-CoV-2.

Furthermore, the clade distribution across the sequences were 64 for the G clade (71.1 %), 14 for GR clade (15.6 %), 6 for GH clade (6.67 %), 4 for GV clade (4.44 %) and 2 for S clade (2.22%), as seen in Fig. 3.

**Figure 3:**
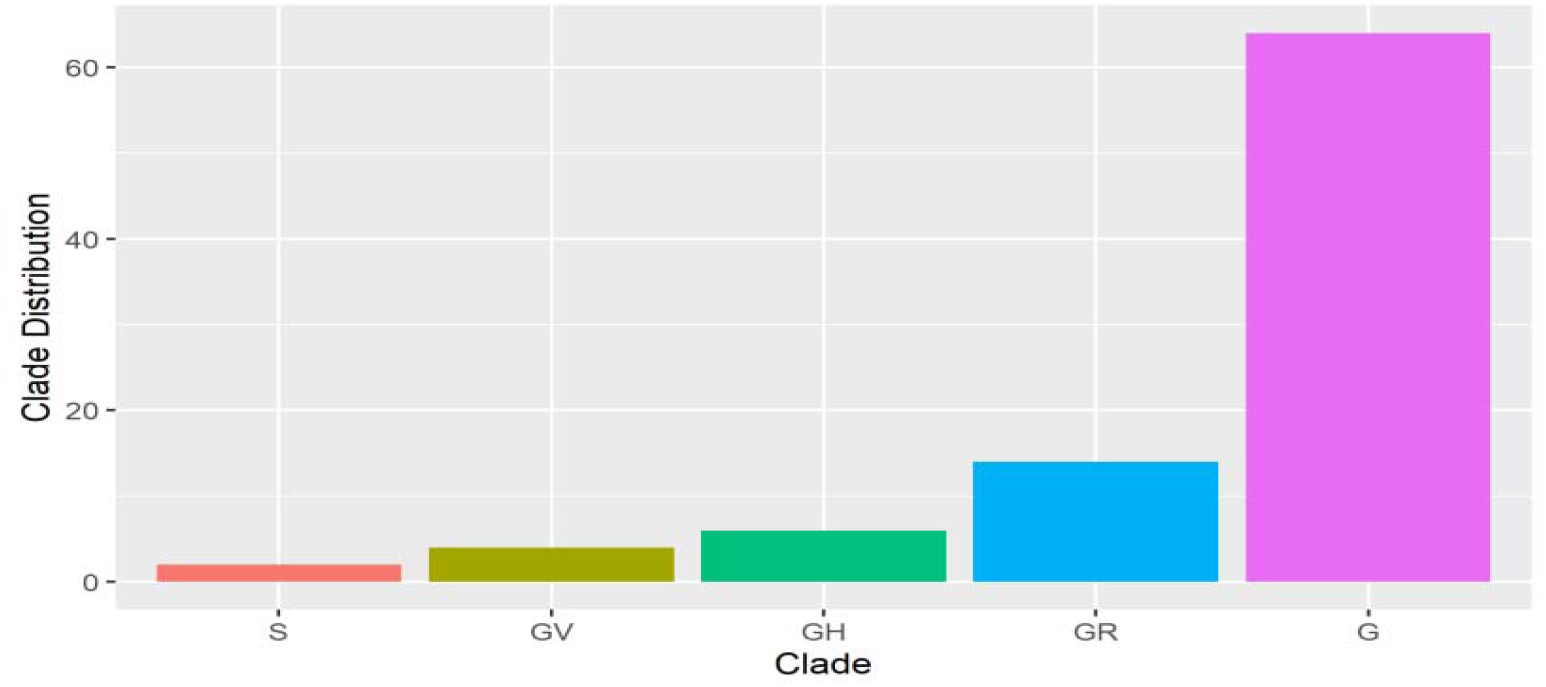
Clade distribution of SARS-CoV-2 from Environmental Source.

Sequel to the Lineage distribution, lineage B.1 has fifty-eight (58), which is the highest number of sequences (64.4 %), followed by lineage B.1.1.119 with twelve (12) sequences (13.3 %). Four (4) Lineages B.1.160, four (4) B.1.177, two (2) B.1.1.33 and two (2) B.1.235 with frequency of (4.44 %), (4.44 %), (2.22 %) and (2.22 %) respectively. The other eight lineages (A.1, A.3, B.1.1.232, B.1.177.24, B.1.2, B.1.258, B.1.283, and B.1.313) occurred once, corresponding to 1.11 % each as seen in Fig. 4.

**Figure 4:**
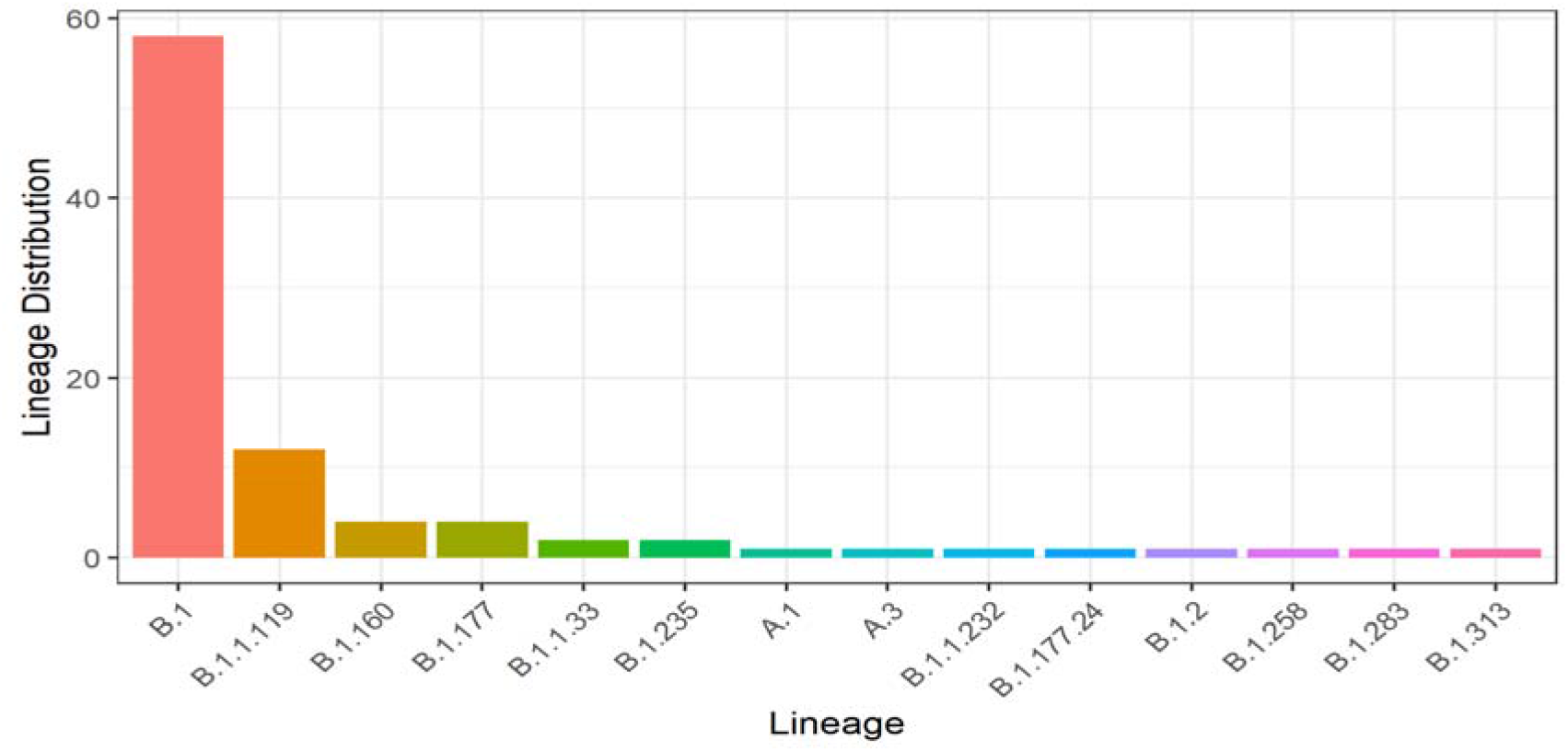
Lineage distribution of SARS-CoV-2 from Environmental Source.

Regarding the specimen obtained relative to their geographical distribution, sixty-eight samples from both Austria (65) and Liechtenstein (3) were isolated from raw sewage Source, while two samples from Brazil were gotten from sewage. Fourteen samples from China were obtained from the environmental swab (5) and the outer package of cold chain products (9), respectively. Six samples from the USA were from the air (2), breathing air (1), door handle (1), environmental swab (1), and wastewater (1), as shown in Fig.5.

**Figure 5:**
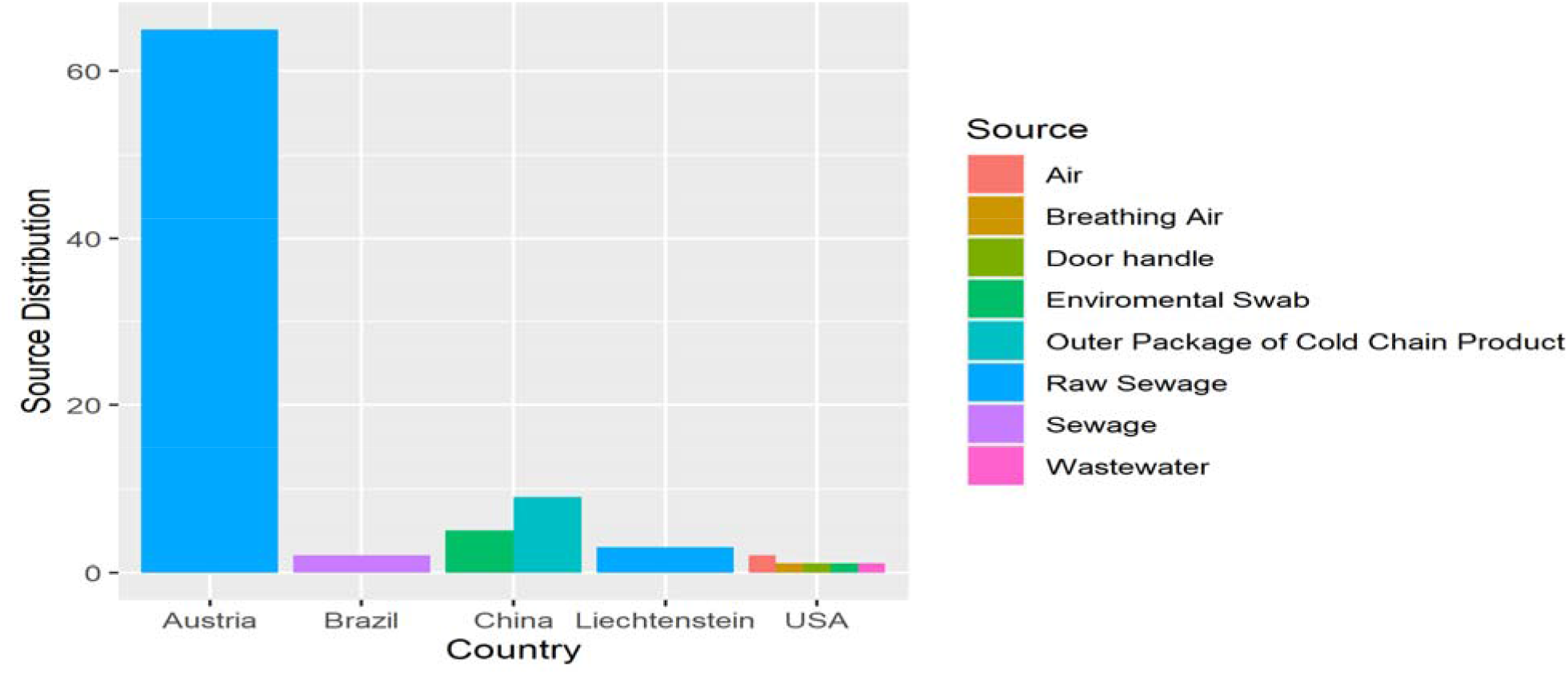
Environmental Source distribution of SARS-CoV-2 per Country.

In addition, the clade obtained relative to the geographical distribution shows four different clades from Austria in the order of G (56), GH (3), GR (2), GV (4), then followed by the USA with three; G (1), GH (3), S (2) and subsequently by China with two; G (4), GR (10), while Brazil and Liechtenstein have GR (2) and G (3) clade respectively as seen in Fig.6.

**Figure 6:**
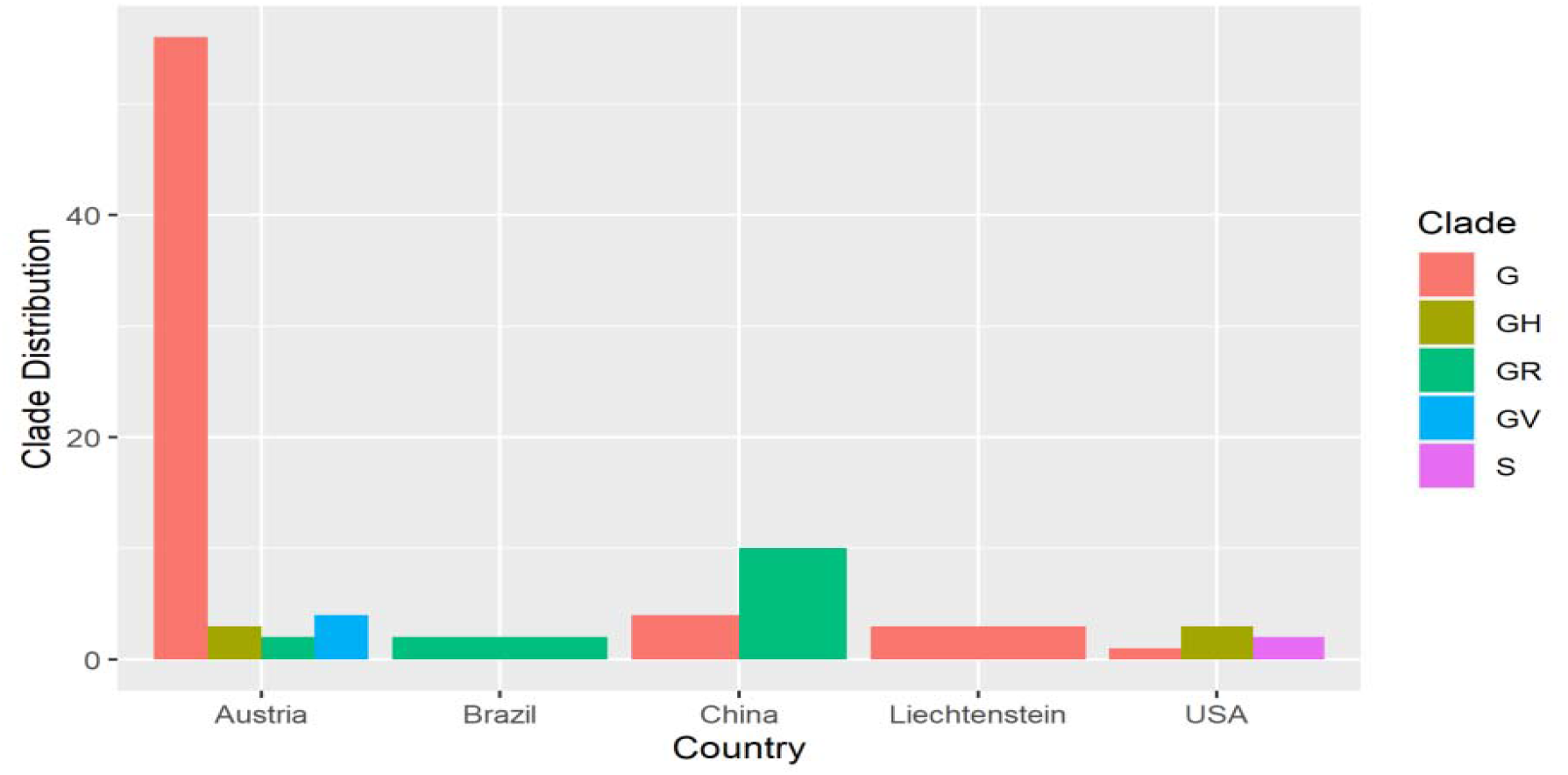
Clade distribution of SARS-CoV-2 from Environmental Source per country.

However, the lineage obtained with respect to the geographical distribution reveals that eight lineages B.1 (50), B.1.1.119 (2), B.1.1.232 (1), B.1.160 (4), B.1.177 (4), B.1.177.24 (1), B.1.235 (2), B.1.258 (1) were originated from Austria, six lineages A.1 (1), A.3 (1), B.1 (1), B.1.2 (1), B.1.283 (1), B.1.313 (1) from USA, two lineages B.1 (4), B.1.1.119 (10) from China. Also, Brazil and Liechtenstein have lineage B.1.133 (2) and B.1 (3), respectively, as seen in Fig.7.

**Figure 7:**
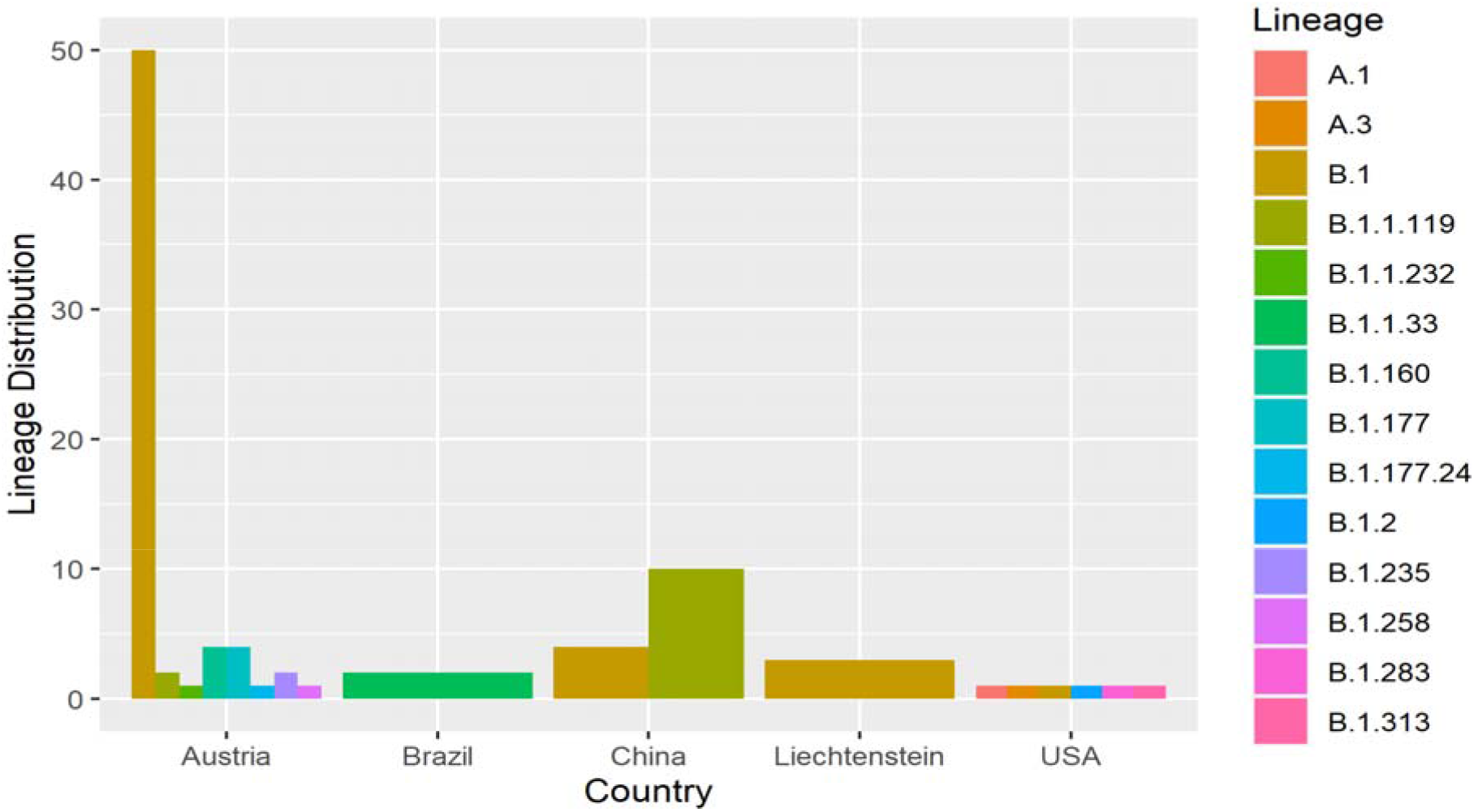
Lineage distribution of SARS-CoV-2 from Environmental Source per Country.

### Multiple Sequence Alignment

The multiple sequence alignment carried out shows a mutation in the genome of the isolated nucleotide sequences, as seen in Fig.8 below. Eighteen (18) of the nucleotide sequences showed a 90% similarity with the Wuhan-Hu-1 reference genome indicating a common source of evolution.

**Figure 8:**
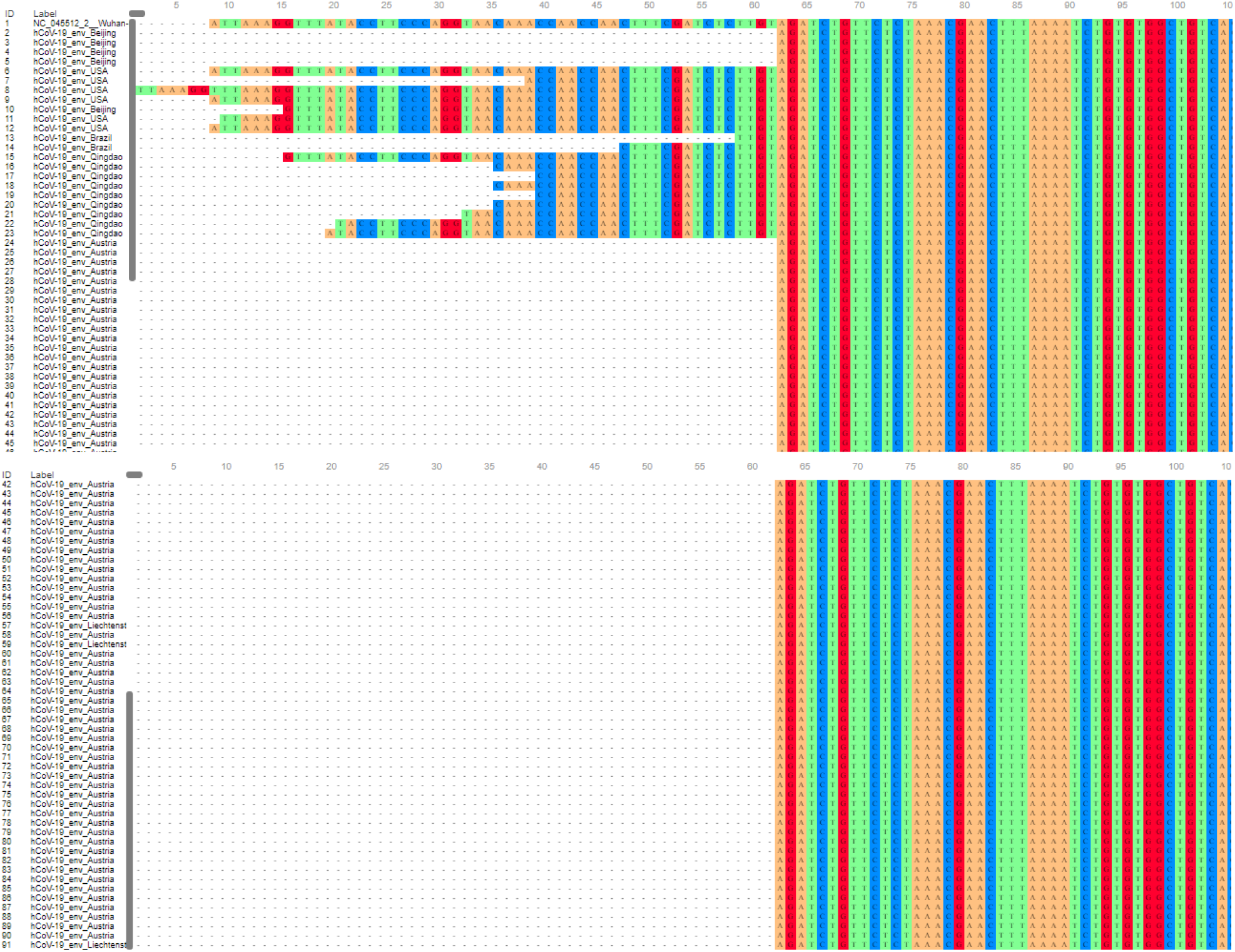
Multiple Sequence Alignment of SARs-CoV2 from Environmental Source.

### Phylogenetic Analysis

Primarily, for the 90 SAR-CoV-2 sequences retrieved, a rooted circular tree was constructed to explain the spread, distribution, and evolutionary process of SARS-CoV-2 in the environment across five (5) countries, as indicated in Fig.9.

**Figure 9:**
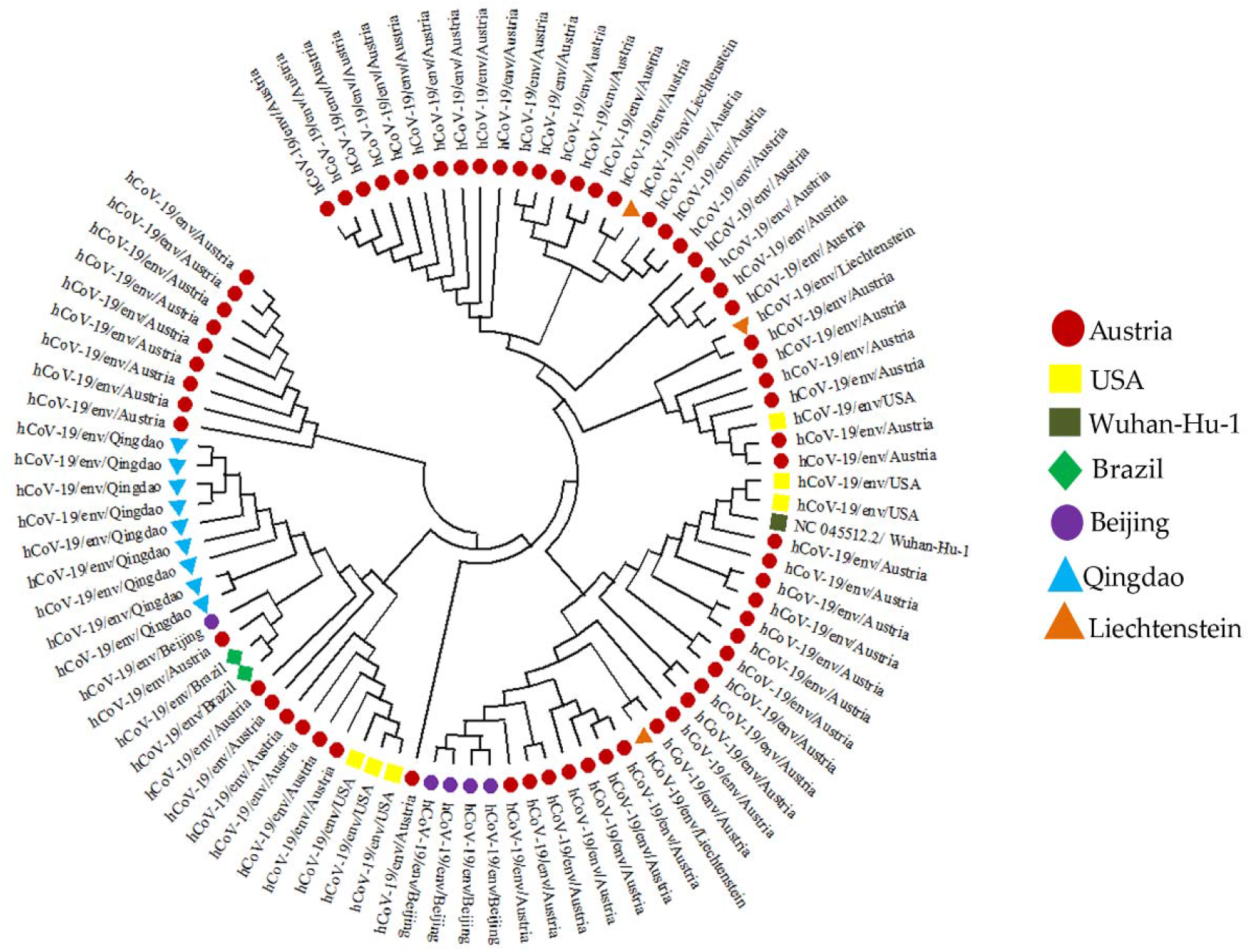
Phylogenetic analysis of SARS-CoV-2 from Environmental Source. (The evolutionary history was inferred using the Maximum Likelihood method and Tamura-Nei model (TNM) (Tamura and Nei, 1993). The bootstrap consensus tree inferred from 50 replicates (Kumar et al., 2018) is taken to represent the evolutionary history of the taxa analyzed (Kumar et al., 2018). Branches corresponding to partitions reproduced in less than 50% bootstrap replicates are collapsed. The tree with the highest log likelihood (−41909.75) is shown. Initial tree(s) for the heuristic search were automatically obtained by applying Neighbor-Join and BioNJ algorithms to a matrix of pairwise distances estimated using the Tamura-Nei model and selecting the topology with a superior log-likelihood value. The tree is drawn to scale, with branch length measured in the sequences. Codon positions included were 1st + 2nd + 3rd + noncoding. This analysis involved 91 nucleotide sequences. There were a total of 29910 positions in the final dataset (Felsenstein, 1985)

## Discussion

The geographic distribution of the SARS-CoV-2 sequence base on their locations revealed that Austria has the highest number of distribution while sequences from Brazil and Liechtenstein have the lowest (Fig. 1). Austria is a country in central Europe with a population of 8.8 million and has a highly developed national epidemiological surveillance program. As of 7 August 2020, contact tracing had been performed for all 21,821 reported SARS-CoV-2–positive cases. Epidemiological clusters were linked to 10,385 of these cases. Integration of phylogenetic analysis of Austrian SARS-CoV-2 sequences with epidemiological data resulted in a strong overlap of these two lines of evidence, with 199 of the 345 sequences (65%) assigned to epidemiological clusters (Popa et al., 2020), in contrast to Brazil and Liechtenstein, where SARS-CoV-2 epidemiological surveillance was insufficient due to their middle-income status. Moreover, there are several controversies about the Brazilian government’s approach to the outbreak. This is evidenced by their President’s resistance to social distancing measures and underestimating the significance of the COVID-19 safety procedure (Lancet, 2020).

Droplets primarily transfer SARS viruses from a respiratory infection at a distance of about 5µm, but they can also be transmitted indirectly through fomite through contact with infected objects, fecal-oral routes, and airborne transmission (Wang et al., 2005; Ye et al., 2016; Kitajima et al., 2020; Naddeo and Liu, 2020). The results of the SARS-CoV-2 environmental source distribution show that specimens isolated from raw sewage have the highest frequency of sequences, while those from breathing air, door handles, and wastewater has the lowest frequency (Figure 2.). SARS-CoV-2 in raw sewage occurrence will always be more than breathing air, door handle because sewage has higher nutrient level to sustain the virus than breathing air, door handle. This relates to Cheng et al. (2020) findings on air and environmental samples of SARS-CoV-2 around hospital patients with COVID-19. Their investigation highlighted that all samples collected from airs source were negative for SARS-CoV-2 RNA in singly isolated patients in airborne infection isolation rooms. Contamination levels on mobile phone surfaces and door handles were found to be lower than in other environmental samples. Following this, Gundy et al. (2020) and Ahmed et al. (2020) reported coronavirus RNA detection in the feces of symptomatic and asymptomatic SARS and COVID-19 patients via raw sewage. They concluded their study by hypothesizing the virus’s persistence in sewage for 14 days at 4°C and 2 days at 20°C, as well as the possibility of detecting its RNA within eight days.

Due to viral susceptibility to disinfectants and poor environmental stability, SARS-CoV-2 may pose a lower risk in wastewater than in sewage (Randazzo et al., 2020; Rimoldi et al., 2020). Reports from two hospital facilities in Beijing, China, reveal that SARS-CoV-2 RNA may be detected in raw sewage, even though there is sometimes no live SARS-CoV-2 following chlorine disinfection. SARS-CoV-2 was isolated from both feces and urine, and its RNA was also found in both (Wang et al., 2019; Sun et al., 2020). SARS-CoV-2, on the other hand, was detected for days on personal items, toilet, room, and floor surfaces, as well as plastic, stainless steel, copper, and cardboard (Santarpia et al., 2020; Van et al., 2020). Persistence on inanimate surfaces such as wood, ceramics, aluminum, glass, waste, containers, bags were also recorded for days. (Santarpia et al., 2020; Van et al., 2020).

The presence of SARS-CoV-2 in sewage and stool could lead to faecal-oral transmission via aerosolization from toilet flushing, a leaky plumbing system, and other sources (Heller et al., 2020). This could be supported by the findings of Van et al. (2020), as observed from this study’s findings. They reported a higher incidence of SARS-CoV-2 viral aerosol transmission in feces among infected individuals in Italian cities, probably attributable to aerosol exposure. In the wide global spread of COVID-19, aerosol transmission could be a more plausible route than faecal-oral transmission. According to findings from previous research, some researchers confirm the spread of SARS-CoV-2 by air as the main route of transmission.

In contrast, other researchers supported aerosol transmission based on their support for asymptomatic individuals (Van et al., 2020). Morawska and Cao (2020) demonstrated that SARS-CoV-2 could remain viable and infectious in aerosols for hours and on surfaces for up to days, highlighting the plausibility of both aerosol and fomite transmission. Furthermore, Fear et al. (2020) observed that SARS-CoV-2 generally maintains infectivity while airborne over short distances and persist for more extended periods than when produced as respiratory particles, based on their findings. In most developing countries with poor waste management strategies, waste collectors and pickers may transfer contaminated solid waste and then retransmitted it back to the community (Nghiem et al., 2020).

Coronaviruses are genetically diverse, with a high proclivity for genetic mutations and gene recombination, which increases the possibility of interspecies transmission (Helmy et al., 2020). According to our findings, Austria has the most significant number of sequences, clades, and lineages contributed to the database. In contrast, sequences from Brazil and Liechtenstein have the lowest number of sequences, clades, and lineages (Figures 5 -7). Eighteen (18) nucleotide sequences from this study’s multiple sequence alignment shared a 90% similarity with the Wuhan-Hu-1 reference genome, indicating common evolutionary origins (Figure 8). This could be linked to the patients’ travel history to the infection’s hotspot (China). The phylodynamic analyses revealed when an epidemic began in a given population and the time of the most recent common ancestor of sampled virus isolates (Sang, 2020). The phylogenetic analysis of SARS-CoV-2 specimen sequences (Fig. 9) indicates that the sequence from the United States has numerous points of interaction with the sequence from Austria.

In contrast, the sequence from China has similar homology with the sequence from Austria, although the sequence from Qingdao is primarily exclusive to Qingdao, the sequences from Brazil have a lot in common with the sequences from Austria and Qingdao. The phylogeny of SARS-CoV-2 whole-genome sequences from Austria, the United States, China, Brazil, and Liechtenstein indicated that the SARS-CoV-2 viruses were all clustered together, irrespective of sequence geographic location. The results obtained suggest the possibility of a recent common ancestor for all SARS-CoV-2 or the transmission of the virus strain across the countries.

In conclusion, this study shows the interconnectedness of the genetic relatedness of SARS-CoV-2 isolates from various geographic locations, all of which are locked in the same cluster regardless of their environmental specimen. These show how they could have shared an ancestor. Furthermore, the high prevalence of SARS-CoV-2 from environmental sources in specimens isolated from raw sewage samples suggests that sewage may be a possible route for virus transmission. To further prevent the spread of the deadly virus, sewage generated from various healthcare and isolation centers must be properly treated before being discharged into the environment.

## Data Availability

GISAID

https://platform.epicov.org/epi3/frontend#1f1ef

## Acknowledgment

The authors are grateful to the GISAID community and authors from the originating laboratories responsible for the genetic sequence data shared via the GISAID Initiative, on which this research is based. Special thanks to Helix Biogen Institute, Ogbomoso, Nigeria, for the technical support.

## Notes

### Competing Interest Statement

The authors have declared no competing interest.

### Funding Statement

No Funding

### Author Declarations

There is no need it is a secondary data obtained from Data GISAID

