## Supplementary file (S1). for "Geographic and Phylodynamic Distribution of SARS-CoV-2 from Environmental Origin"

Virus name	Accession ID	Collection date	Location	Host	Additional location information	Gender	Patient age	Patient status	Passage	Specimen	Additional host information	Lineage	Clade

hCoV-19/env/Wuhan/IVDC-HBF13-20/2020	EPI_ISL_408514	1/1/2020	Asia / China / Hubei / Wuhan	Environment Huanan Seafood Market	unknown	n/a	unknown	P1	B	L

hCoV-19/env/Wuhan/IVDC-HBF13-21/2020	EPI_ISL_408515	1/1/2020	Asia / China / Hubei / Wuhan	Environment Huanan Seafood Market	unknown	n/a	unknown	P1	B	L

hCoV-19/env/Beijing/BJ2452/2020	EPI_ISL_430743	3/14/2020	Asia / China / Beijing	Environment	unknown	n/a	unknown	Original	Enviromental swab	B.1	G

hCoV-19/env/Beijing/BJ2454/2020	EPI_ISL_430744	3/14/2020	Asia / China / Beijing	Environment	unknown	n/a	unknown	Original	Enviromental swab	B.1	G

hCoV-19/env/Beijing/BJ2456/2020	EPI_ISL_430745	3/14/2020	Asia / China / Beijing	Environment	unknown	n/a	unknown	Original	Enviromental swab	B.1	G

hCoV-19/env/Beijing/BJ2460/2020	EPI_ISL_430746	3/14/2020	Asia / China / Beijing	Environment	unknown	n/a	unknown	Original	Enviromental swab	B.1	G

hCoV-19/env/USA/FL-UF3/2020	EPI_ISL_434677	3/25/2020	North America / USA / Florida	Environment Breathing air using VIVAs air sampler	unknown	n/a	unknown	Original	Breathing air using VIVAs Air sampler	A.3	S

hCoV-19/env/USA/MT-BWRF-0327/2020	EPI_ISL_437434	3/27/2020	North America / USA / Montana	Environment Bozeman Water Reclamation Facility	unknown	unknown	unknown	Original	Wastewater	Wastewater	B.1.283	GH

hCoV-19/env/USA/FL-UF11/2020	EPI_ISL_447900	2/21/2020	North America / USA / Florida	Environment	unknown	unknown	unknown	Original	Door handle	A.1	S

hCoV-19/env/USA/UN-UF-12/2020	EPI_ISL_455682	4/23/2020	North America / USA / Florida	Environment	unknown	unknown	unknown	Original	Air	B.1.2	GH

hCoV-19/env/Beijing/IVDC-03-06/2020	EPI_ISL_469256	6/11/2020	Asia / China / Beijing	Environment Xinfadi Wholesale Market	unknown	unknown	unknown	Original	Enviromental swab	B.1.1.119	GR

hCoV-19/env/USA/FL-UF-20/2020	EPI_ISL_477160	4/13/2020	North America / USA / Florida	Environment Air in the hospital room of COVID-19 patients	unknown	unknown	unknown	Vero E6	Air sample	B.1.313	GH

hCoV-19/env/USA/UT-CDC-1251/2020	EPI_ISL_510160	4/25/2020	North America / USA / Utah	Environment	unknown	unknown	unknown	Original	swab	B.1	G

hCoV-19/env/USA/NV-NSPHL-A0128/2020	EPI_ISL_515396	4/29/2020	North America / USA / Nevada	Environment	unknown	unknown	unknown	Original	B.23	L

hCoV-19/env/USA/NV-NSPHL-A0130/2020	EPI_ISL_515398	4/29/2020	North America / USA / Nevada	Environment	unknown	unknown	unknown	Original	B.1.1.64	O

hCoV-19/env/Netherlands/UT-92848-N/2020	EPI_ISL_539306	4/1/2020	Europe / Netherlands / Utrecht / Amersfoort	Environment	unknown	unknown	unknown	Original	Wastewater	B.1	O

hCoV-19/env/Netherlands/NH-92508-N/2020	EPI_ISL_539307	3/25/2020	Europe / Netherlands / North Holland / Amsterdam West	Environment	unknown	unknown	unknown	Original	Wastewater	B.1	O

hCoV-19/env/Netherlands/NH-92852-N1/202	EPI_ISL_539308	4/1/2020	Europe / Netherlands / North Holland / Amsterdam West	Environment	unknown	unknown	unknown	Original	Wastewater	B.1	G

hCoV-19/env/Netherlands/NH-92852-N2/202	EPI_ISL_539309	4/1/2020	Europe / Netherlands / North Holland / Amsterdam West	Environment	unknown	unknown	unknown	Original	Wastewater	B.1	G

hCoV-19/env/Netherlands/FR-92719-N1/2020	EPI_ISL_539314	3/30/2020	Europe / Netherlands / Friesland / Franeker	Environment	unknown	unknown	unknown	Original	Wastewater	B.1.1.292	GR

hCoV-19/env/Netherlands/NH-92723-N/2020	EPI_ISL_539325	3/30/2020	Europe / Netherlands / North Holland / Wervershoof	Environment	unknown	unknown	unknown	Original	Wastewater	B.1.260	O

hCoV-19/env/Brazil/RJ-4712/2020	EPI_ISL_541397	4/22/2020	South America / Brazil / Rio de Janeiro / Niteroi / Boa Esperanca	Environment	unknown	unknown	unknown	Original	Sewage	B.1.1.33	GR

hCoV-19/env/Brazil/RJ-4830/2020	EPI_ISL_541399	6/2/2020	South America / Brazil / Rio de Janeiro / Niteroi / Icarai	Environment	unknown	unknown	unknown	Original	Sewage	B.1.1.33	GR

hCoV-19/env/Qingdao/IVDC-03-10/2020	EPI_ISL_591272	9/24/2020	Asia / China / Shandong / Qingdao	Environment	NA	n/a	NA	Original	Outer packaging of cold chain products	B.1.1.119	GR

hCoV-19/env/Qingdao/IVDC-04-10/2020	EPI_ISL_591273	9/24/2020	Asia / China / Shandong / Qingdao	Environment	NA	n/a	NA	Original	Outer packaging of cold chain products	B.1.1.119	GR

hCoV-19/env/Qingdao/IVDC-05-10/2020	EPI_ISL_591274	9/24/2020	Asia / China / Shandong / Qingdao	Environment	NA	n/a	NA	Original	Outer packaging of cold chain products	B.1.1.119	GR

hCoV-19/env/Qingdao/IVDC-06-10/2020	EPI_ISL_591275	9/24/2020	Asia / China / Shandong / Qingdao	Environment	NA	n/a	NA	Original	Outer packaging of cold chain products	B.1.1.119	GR

hCoV-19/env/Qingdao/IVDC-07-10/2020	EPI_ISL_591276	9/24/2020	Asia / China / Shandong / Qingdao	Environment	NA	n/a	NA	Original	Outer packaging of cold chain products	B.1.1.119	GR

hCoV-19/env/Qingdao/IVDC-08-10/2020	EPI_ISL_591277	9/24/2020	Asia / China / Shandong / Qingdao	Environment	NA	n/a	NA	Original	Outer packaging of cold chain products	B.1.1.119	GR

hCoV-19/env/Qingdao/IVDC-09-10/2020	EPI_ISL_591278	9/24/2020	Asia / China / Shandong / Qingdao	Environment	NA	n/a	NA	Original	Outer packaging of cold chain products	B.1.1.119	GR

hCoV-19/env/Qingdao/IVDC-010-10/2020	EPI_ISL_591279	9/27/2020	Asia / China / Shandong / Qingdao	Environment	NA	n/a	NA	Original	Outer packaging of cold chain products	B.1.1.119	GR

hCoV-19/env/Qingdao/IVDC-011-10/2020	EPI_ISL_591280	10/7/2020	Asia / China / Shandong / Qingdao	Environment	NA	n/a	NA	Vero C2	Outer packaging of cold chain products isolated from Vero cells	B.1.1.119	GR

hCoV-19/env/Hong Kong/VN20001008/2020	EPI_ISL_733568	12/10/2020	Asia / Hong Kong	Environment	NA	NA	NA	Original	B.1.36.27	GH hCoV-19/env/Austria/CeMM1446/2020	EPI_ISL_853718	10/19/2020	Europe / Austria / Salzburg	Environment Sewage treatment plant for Bischofshofen, Goldegg, Kleinarl, Pfarrwerfen, Sankt Johann im Pongau, Sankt Veit im Pongau, Schwarzach im Pongau, Wagrain, W unknown	unknown	unknown	Original	Raw Sewage, PEG Precipitation	B.1.1.119	G hCoV-19/env/Austria/CeMM1465/2020	EPI_ISL_853719	10/25/2020	Europe / Austria / Salzburg	Environment Sewage treatment plant for Salzburg, Anif, Anthering, Bergheim, Elixhausen, Elsbethen, Eugendorf, Grödig, Hallwang, Koppl, Puch bei Hallein, Wals-Siezenheim unknown	unknown	unknown	Original	Raw Sewage, PEG Precipitation	B.1	G hCoV-19/env/Austria/CeMM1452/2020	EPI_ISL_853720	10/18/2020	Europe / Austria / Vorarlberg	Environment Sewage treatment plant for Bregenz, Kennelbach, Lochau	unknown	unknown	unknown	Original	Raw Sewage, PEG Precipitation	B.1.177	GV hCoV-19/env/Austria/CeMM2219/2020	EPI_ISL_853722	12/28/2020	Europe / Austria / Salzburg	Environment Sewage treatment plant for Leogang, Maishofen, Maria Alm am Steinernen Meer, Saalfelden am Steinernen Meer, Viehhofen	unknown	unknown	unknown	Original	Raw Sewage, PEG Precipitation	B.1	G hCoV-19/env/Austria/CeMM2022/2020	EPI_ISL_853723	11/29/2020	Europe / Austria / Vienna	Environment	unknown	unknown	unknown	Original	Raw Sewage, PEG Precipitation	B.1	GH hCoV-19/env/Austria/CeMM2224/2020	EPI_ISL_853724	12/20/2020	Europe / Austria / Salzburg	Environment	Sewage treatment plant for Salzburg, Anif, Anthering, Bergheim, Elixhausen, Elsbethen, Eugendorf, Grödig, Hallwang, Koppl, Puch bei Hallein, Wals-Siezenheim unknown	unknown	unknown	Original	Raw Sewage, PEG Precipitation	B.1	G hCoV-19/env/Austria/CeMM2232/2020	EPI_ISL_853725	12/14/2020	Europe / Austria / Carinthia	Environment	Sewage treatment plant for Hohenems, Altach, Götzis, Koblach, Mäder	unknown	unknown	unknown	Original	Raw Sewage, PEG Precipitation	B.1	G hCoV-19/env/Austria/CeMM2233/2020	EPI_ISL_853726	12/20/2020	Europe / Austria / Carinthia	Environment	Sewage treatment plant for Hohenems, Altach, Götzis, Koblach, Mäder	unknown	unknown	unknown	Original	Raw Sewage, PEG Precipitation	B.1	G hCoV-19/env/Austria/CeMM2234/2021	EPI_ISL_853727	1/2/2021	Europe / Austria / Carinthia	Environment	Sewage treatment plant for Hohenems, Altach, Götzis, Koblach, Mäder	unknown	unknown	unknown	Original	Raw Sewage, PEG Precipitation	B.1	G hCoV-19/env/Austria/CeMM1448/2020	EPI_ISL_853728	10/29/2020	Europe / Austria / Salzburg	Environment	Sewage treatment plant for Golling an der Salzach, Kuchl, Sankt Koloman, Scheffau am Tennengebirge	unknown	unknown	unknown	Original	Raw Sewage, PEG Precipitation	B.1.160	GH hCoV-19/env/Austria/CeMM1447/2020	EPI_ISL_853732	10/18/2020	Europe / Austria / Salzburg	Environment	Sewage treatment plant for Golling an der Salzach, Kuchl, Sankt Koloman, Scheffau am Tennengebirge	unknown	unknown	unknown	Original	Raw Sewage, PEG Precipitation	B.1.160	G hCoV-19/env/Austria/CeMM2210/2020	EPI_ISL_853739	12/15/2020	Europe / Austria / Carinthia	Environment	Sewage treatment plant for Velden am Wörther See, Wernberg, St. Jakob im Rosental	unknown	unknown	unknown	Original	Raw Sewage, PEG Precipitation	B.1	G hCoV-19/env/Austria/CeMM2226/2020	EPI_ISL_853740	12/12/2020	Europe / Austria / Salzburg	Environment	Sewage treatment plant for Golling an der Salzach, Kuchl, Sankt Koloman, Scheffau am Tennengebirge	unknown	unknown	unknown	Original	Raw Sewage, PEG Precipitation	B.1.160	G hCoV-19/env/Austria/CeMM2211/2020	EPI_ISL_853742	12/29/2020	Europe / Austria / Carinthia	Environment	Sewage treatment plant for Villach	unknown	unknown	unknown	Original	Raw Sewage, PEG Precipitation	B.1	G hCoV-19/env/Austria/CeMM2225/2020	EPI_ISL_853743	12/27/2020	Europe / Austria / Salzburg	Environment	Sewage treatment plant for Salzburg, Anif, Anthering, Bergheim, Elixhausen, Elsbethen, Eugendorf, Grödig, Hallwang, Koppl, Puch bei Hallein, Wals-Siezenheim unknown	unknown	unknown	Original	Raw Sewage, PEG Precipitation	B.1	G hCoV-19/env/Austria/CeMM2229/2020	EPI_ISL_853744	12/15/2020	Europe / Austria / Salzburg	Environment	Sewage treatment plant for Kaprun, Zell am See, Maishofen, Piesendorf	unknown	unknown	unknown	Original	Raw Sewage, PEG Precipitation	B.1	G hCoV-19/env/Austria/CeMM2230/2020	EPI_ISL_853745	12/27/2020	Europe / Austria / Salzburg	Environment	Sewage treatment plant for Kaprun, Zell am See, Maishofen, Piesendorf	unknown	unknown	unknown	Original	Raw Sewage, PEG Precipitation	B.1.177	G hCoV-19/env/Austria/CeMM2218/2020	EPI_ISL_853747	12/15/2020	Europe / Austria / Salzburg	Environment	Sewage treatment plant for Filzmoos, Eben im Pongau, Hüttau, Sankt Martin am Tennengebirge	unknown	unknown	unknown	Original	Raw Sewage, PEG Precipitation	B.1	G hCoV-19/env/Austria/CeMM2222/2021	EPI_ISL_853748	1/3/2021	Europe / Austria / Salzburg	Environment	Sewage treatment plant for Bischofshofen, Goldegg, Kleinarl, Pfarrwerfen, Sankt Johann im Pongau, Sankt Veit im Pongau, Schwarzach im Pongau, Wagrain, W unknown	unknown	unknown	Original	Raw Sewage, PEG Precipitation	B.1	G hCoV-19/env/Austria/CeMM2212/2020	EPI_ISL_853750	12/17/2020	Europe / Austria / Carinthia	Environment	Sewage treatment plant for Klagenfurt, Ebenthal, Maria Saal, Moosburg, Schiefling, Keutschach, Köttmannsdorf, Krumpendorf, Maria Rain, Maria Wörth, Pörts unknown	unknown	unknown	Original	Raw Sewage, PEG Precipitation	B.1	G hCoV-19/env/Austria/CeMM2019/2020	EPI_ISL_853752	11/2/2020	Europe / Austria / Vienna	Environment	unknown	unknown	unknown	Original	Raw Sewage, PEG Precipitation	B.1.1.119	GR hCoV-19/env/Austria/CeMM1442/2020	EPI_ISL_853755	10/18/2020	Europe / Austria / Salzburg	Environment	Sewage treatment plant for Salzburg, Anif, Anthering, Bergheim, Elixhausen, Elsbethen, Eugendorf, Grödig, Hallwang, Koppl, Puch bei Hallein, Wals-Siezenheim unknown	unknown	unknown	Original	Raw Sewage, PEG Precipitation	B.1	G hCoV-19/env/Austria/CeMM2214/2020	EPI_ISL_853756	12/16/2020	Europe / Austria / Styria	Environment	Sewage treatment plant for Stadl-Predlitz, Moosburg, Steindorf am Ossiacher See, Steuerberg, Feldkirchen in Kärnten, Gnesau, Himmelberg, Reichenau, St. Ur	unknown	unknown	unknown	Original	Raw Sewage, PEG Precipitation	B.1	G hCoV-19/env/Austria/CeMM2228/2021	EPI_ISL_853760	1/3/2021	Europe / Austria / Salzburg	Environment	Sewage treatment plant for Golling an der Salzach, Kuchl, Sankt Koloman, Scheffau am Tennengebirge	unknown	unknown	unknown	Original	Raw Sewage, PEG Precipitation	B.1	G hCoV-19/env/Austria/CeMM1456/2020	EPI_ISL_853761	10/19/2020	Europe / Austria / Carinthia	Environment Sewage treatment plant for Villach	unknown	unknown	unknown	Original	Raw Sewage, PEG Precipitation	B.1	G

hCoV-19/env/Austria/CeMM2020/2020	EPI_ISL_853762	11/8/2020	Europe / Austria / Vienna	Environment	unknown	unknown	unknown	Original	Raw Sewage, PEG Precipitation	B.1	G hCoV-19/env/Austria/CeMM2220/2020	EPI_ISL_853763	12/18/2020	Europe / Austria / Salzburg	Environment Sewage treatment plant for Bischofshofen, Goldegg, Kleinarl, Pfarrwerfen, Sankt Johann im Pongau, Sankt Veit im Pongau, Schwarzach im Pongau, Wagrain, W unknown	unknown	unknown	Original	Raw Sewage, PEG Precipitation	B.1	G hCoV-19/env/Austria/CeMM2223/2020	EPI_ISL_853765	12/15/2020	Europe / Austria / Salzburg	Environment Sewage treatment plant for Altenmarkt im Pongau, Flachau, Radstadt, Untertauern, Tweng	unknown	unknown	unknown	Original	Raw Sewage, PEG Precipitation	B.1	G hCoV-19/env/Austria/CeMM1954/2020	EPI_ISL_853767	11/18/2020	Europe / Austria / Salzburg	Environment Sewage treatment plant for Salzburg, Anif, Anthering, Bergheim, Elixhausen, Elsbethen, Eugendorf, Grödig, Hallwang, Koppl, Puch bei Hallein, Wals-Siezenheim unknown	unknown	unknown	Original	Raw Sewage, PEG Precipitation	B.1	G hCoV-19/env/Austria/CeMM1462/2020	EPI_ISL_853783	10/16/2020	Europe / Austria / Salzburg	Environment Sewage treatment plant for Salzburg, Anif, Anthering, Bergheim, Elixhausen, Elsbethen, Eugendorf, Grödig, Hallwang, Koppl, Puch bei Hallein, Wals-Siezenheim unknown	unknown	unknown	Original	Raw Sewage, PEG Precipitation	B.1	G hCoV-19/env/Austria/CeMM1399/2020	EPI_ISL_853785	8/31/2020	Europe / Austria / Carinthia	Environment Sewage treatment plant for Wernberg, Velden, St. Jakob im Rosental, Rosegg	unknown	unknown	unknown	Original	Raw Sewage, PEG Precipitation	B.1.1.232	GR hCoV-19/env/Austria/CeMM2231/2020	EPI_ISL_853786	12/16/2020	Europe / Austria / Vorarlberg	Environment Sewage treatment plant for Bregenz, Kennelbach, Lochau	unknown	unknown	unknown	Original	Raw Sewage, PEG Precipitation	B.1	G

hCoV-19/env/Austria/CeMM2021/2020	EPI_ISL_853787	11/15/2020	Europe / Austria / Vienna	Environment	unknown	unknown	unknown	Original	Raw Sewage, PEG Precipitation	B.1	GH

hCoV-19/env/Liechtenstein/CeMM2216/2020 EPI_ISL_853788	12/23/2020	Europe / Liechtenstein	Environment	unknown	unknown	unknown	Original	Raw Sewage, PEG Precipitation	B.1	G hCoV-19/env/Austria/CeMM1978/2020	EPI_ISL_853792	11/17/2020	Europe / Austria / Salzburg	Environment Sewage treatment plant for Golling an der Salzach, Kuchl, Sankt Koloman, Scheffau am Tennengebirge	unknown	unknown	unknown	Original	Raw Sewage, PEG Precipitation	B.1	G hCoV-19/env/Liechtenstein/CeMM2215/2020 EPI_ISL_853796	12/20/2020	Europe / Liechtenstein	Environment	unknown	unknown	unknown	Original	Raw Sewage, PEG Precipitation	B.1	G hCoV-19/env/Austria/CeMM1975/2020	EPI_ISL_853802	12/4/2020	Europe / Austria / Salzburg	Environment Sewage treatment plant for Altenmarkt im Pongau, Flachau, Radstadt, Untertauern, Tweng	unknown	unknown	unknown	Original	Raw Sewage, PEG Precipitation	B.1	G hCoV-19/env/Austria/CeMM2227/2020	EPI_ISL_853806	12/24/2020	Europe / Austria / Salzburg	Environment Sewage treatment plant for Golling an der Salzach, Kuchl, Sankt Koloman, Scheffau am Tennengebirge	unknown	unknown	unknown	Original	Raw Sewage, PEG Precipitation	B.1.160	G hCoV-19/env/Austria/CeMM1963/2020	EPI_ISL_853814	12/2/2020	Europe / Austria / Carinthia	Environment Sewage treatment plant for Hohenems, Altach, Götzis, Koblach, Mäder	unknown	unknown	unknown	Original	Raw Sewage, PEG Precipitation	B.1.235	G hCoV-19/env/Austria/CeMM2018/2020	EPI_ISL_853816	10/25/2020	Europe / Austria / Vienna	Environment	unknown	unknown	unknown	Original	Raw Sewage, PEG Precipitation	B.1	G hCoV-19/env/Austria/CeMM1953/2020	EPI_ISL_853929	11/6/2020	Europe / Austria / Salzburg	Environment	Sewage treatment plant for Salzburg, Anif, Anthering, Bergheim, Elixhausen, Elsbethen, Eugendorf, Grödig, Hallwang, Koppl, Puch bei Hallein, Wals-Siezenheim unknown	unknown	unknown	Original	Raw Sewage, PEG Precipitation	B.1	G hCoV-19/env/Austria/CeMM1955/2020	EPI_ISL_853930	11/30/2020	Europe / Austria / Salzburg	Environment	Sewage treatment plant for Salzburg, Anif, Anthering, Bergheim, Elixhausen, Elsbethen, Eugendorf, Grödig, Hallwang, Koppl, Puch bei Hallein, Wals-Siezenheim unknown	unknown	unknown	Original	Raw Sewage, PEG Precipitation	B.1	G hCoV-19/env/Austria/CeMM1956/2020	EPI_ISL_853931	12/8/2020	Europe / Austria / Salzburg	Environment	Sewage treatment plant for Salzburg, Anif, Anthering, Bergheim, Elixhausen, Elsbethen, Eugendorf, Grödig, Hallwang, Koppl, Puch bei Hallein, Wals-Siezenheim unknown	unknown	unknown	Original	Raw Sewage, PEG Precipitation	B.1	G hCoV-19/env/Austria/CeMM1957/2020	EPI_ISL_853932	11/4/2020	Europe / Austria / Carinthia	Environment	Sewage treatment plant for Velden am Wörther See, Wernberg, St. Jakob im Rosental	unknown	unknown	unknown	Original	Raw Sewage, PEG Precipitation	B.1	G hCoV-19/env/Austria/CeMM1958/2020	EPI_ISL_853933	11/18/2020	Europe / Austria / Carinthia	Environment	Sewage treatment plant for Velden am Wörther See, Wernberg, St. Jakob im Rosental	unknown	unknown	unknown	Original	Raw Sewage, PEG Precipitation	B.1	G hCoV-19/env/Austria/CeMM1959/2020	EPI_ISL_853934	12/2/2020	Europe / Austria / Carinthia	Environment	Sewage treatment plant for Velden am Wörther See, Wernberg, St. Jakob im Rosental	unknown	unknown	unknown	Original	Raw Sewage, PEG Precipitation	B.1	G hCoV-19/env/Austria/CeMM1960/2020	EPI_ISL_853935	12/13/2020	Europe / Austria / Carinthia	Environment	Sewage treatment plant for Velden am Wörther See, Wernberg, St. Jakob im Rosental	unknown	unknown	unknown	Original	Raw Sewage, PEG Precipitation	B.1	G hCoV-19/env/Austria/CeMM1961/2020	EPI_ISL_853936	11/1/2020	Europe / Austria / Carinthia	Environment	Sewage treatment plant for Hohenems, Altach, Götzis, Koblach, Mäder	unknown	unknown	unknown	Original	Raw Sewage, PEG Precipitation	B.1.177	G hCoV-19/env/Austria/CeMM1962/2020	EPI_ISL_853937	11/15/2020	Europe / Austria / Carinthia	Environment	Sewage treatment plant for Hohenems, Altach, Götzis, Koblach, Mäder	unknown	unknown	unknown	Original	Raw Sewage, PEG Precipitation	B.1	G hCoV-19/env/Austria/CeMM1964/2020	EPI_ISL_853938	11/1/2020	Europe / Austria / Vorarlberg	Environment	Sewage treatment plant for Bregenz, Kennelbach, Lochau	unknown	unknown	unknown	Original	Raw Sewage, PEG Precipitation	B.1.177.24	GV

hCoV-19/env/Austria/CeMM1965/2020	EPI_ISL_853939	11/15/2020	Europe / Austria / Vorarlberg	Environment Sewage treatment plant for Bregenz, Kennelbach, Lochau	unknown	unknown	unknown	Original	Raw Sewage, PEG Precipitation	B.1	G

hCoV-19/env/Austria/CeMM1966/2020	EPI_ISL_853940	12/2/2020	Europe / Austria / Vorarlberg	Environment Sewage treatment plant for Bregenz, Kennelbach, Lochau	unknown	unknown	unknown	Original	Raw Sewage, PEG Precipitation	B.1.177	GV

hCoV-19/env/Austria/CeMM1967/2020	EPI_ISL_853941	11/2/2020	Europe / Austria / Carinthia	Environment Sewage treatment plant for Villach	unknown	unknown	unknown	Original	Raw Sewage, PEG Precipitation	B.1	G

hCoV-19/env/Austria/CeMM1968/2020	EPI_ISL_853942	11/16/2020	Europe / Austria / Carinthia	Environment Sewage treatment plant for Villach	unknown	unknown	unknown	Original	Raw Sewage, PEG Precipitation	B.1	G

hCoV-19/env/Austria/CeMM1969/2020	EPI_ISL_853943	12/3/2020	Europe / Austria / Carinthia	Environment Sewage treatment plant for Villach	unknown	unknown	unknown	Original	Raw Sewage, PEG Precipitation	B.1	G hCoV-19/env/Austria/CeMM1970/2020	EPI_ISL_853944	11/1/2020	Europe / Austria / Salzburg	Environment Sewage treatment plant for Kaprun, Zell am See, Maishofen, Piesendorf	unknown	unknown	unknown	Original	Raw Sewage, PEG Precipitation	B.1	G hCoV-19/env/Austria/CeMM1971/2020	EPI_ISL_853945	11/19/2020	Europe / Austria / Salzburg	Environment Sewage treatment plant for Kaprun, Zell am See, Maishofen, Piesendorf	unknown	unknown	unknown	Original	Raw Sewage, PEG Precipitation	B.1	G hCoV-19/env/Austria/CeMM1972/2020	EPI_ISL_853946	11/5/2020	Europe / Austria / Salzburg	Environment Sewage treatment plant for Kaprun, Zell am See, Maishofen, Piesendorf	unknown	unknown	unknown	Original	Raw Sewage, PEG Precipitation	B.1	G hCoV-19/env/Austria/CeMM1973/2020	EPI_ISL_853947	11/4/2020	Europe / Austria / Salzburg	Environment Sewage treatment plant for Altenmarkt im Pongau, Flachau, Radstadt, Untertauern, Tweng	unknown	unknown	unknown	Original	Raw Sewage, PEG Precipitation	B.1	G hCoV-19/env/Austria/CeMM1974/2020	EPI_ISL_853948	11/20/2020	Europe / Austria / Salzburg	Environment Sewage treatment plant for Altenmarkt im Pongau, Flachau, Radstadt, Untertauern, Tweng	unknown	unknown	unknown	Original	Raw Sewage, PEG Precipitation	B.1	G hCoV-19/env/Austria/CeMM1976/2020	EPI_ISL_853949	11/15/2020	Europe / Austria / Vorarlberg	Environment	Sewage treatment plant for Bildstein, Fußach, Gaißau, Hard, Höchst, Lauterach, Wolfurt, Lustenau	unknown	unknown	unknown	Original	Raw Sewage, PEG Precipitation	B.1	G hCoV-19/env/Austria/CeMM1977/2020	EPI_ISL_853950	12/6/2020	Europe / Austria / Vorarlberg	Environment	Sewage treatment plant for Bildstein, Fußach, Gaißau, Hard, Höchst, Lauterach, Wolfurt, Lustenau	unknown	unknown	unknown	Original	Raw Sewage, PEG Precipitation	B.1.235	GV hCoV-19/env/Austria/CeMM1979/2020	EPI_ISL_853951	12/6/2020	Europe / Austria / Salzburg	Environment	Sewage treatment plant for Golling an der Salzach, Kuchl, Sankt Koloman, Scheffau am Tennengebirge	unknown	unknown	unknown	Original	Raw Sewage, PEG Precipitation	B.1	G hCoV-19/env/Austria/CeMM1980/2020	EPI_ISL_853952	11/15/2020	Europe / Austria / Carinthia	Environment Sewage treatment plant for Klagenfurt am Wörthersee	unknown	unknown	unknown	Original	Raw Sewage, PEG Precipitation	B.1	G

hCoV-19/env/Austria/CeMM1981/2020	EPI_ISL_853953	11/29/2020	Europe / Austria / Carinthia	Environment Sewage treatment plant for Klagenfurt am Wörthersee	unknown	unknown	unknown	Original	Raw Sewage, PEG Precipitation	B.1	G

hCoV-19/env/Austria/CeMM1982/2020	EPI_ISL_853954	12/4/2020	Europe / Austria / Carinthia	Environment Sewage treatment plant for Klagenfurt am Wörthersee	unknown	unknown	unknown	Original	Raw Sewage, PEG Precipitation	B.1	G hCoV-19/env/Austria/CeMM2213/2020	EPI_ISL_853955	12/7/2020	Europe / Austria / Styria	Environment Sewage treatment plant for Stadl-Predlitz, Moosburg, Steindorf am Ossiacher See, Steuerberg, Feldkirchen in Kärnten, Gnesau, Himmelberg, Reichenau, St. Ur unknown	unknown	unknown	Original	Raw Sewage, PEG Precipitation	B.1.258	G hCoV-19/env/Liechtenstein/CeMM2217/2020 EPI_ISL_853956	12/27/2020	Europe / Liechtenstein	Environment	unknown	unknown	unknown	Original	Raw Sewage, PEG Precipitation	B.1	G
